## Supplementary material for "Prognostic accuracy of qSOFA score, SIRS criteria, and EWSs for in-hospital mortality among adult patients presenting with suspected infection to the emergency department (PASSEM): protocol for an international multicentre prospective external validation cohort study": Case report form

Study ID: ____________________

### Screening

#### Eligibility screening

**1.1. Date of birth (Gregorian):** __ /__ /____ (DD/MM/YYYY)

Exclude if age <18 years

**1.2. Sex**

- ☐ Men (1)
- ☐ Women (2)

**1.3. Pregnant** ☐ Yes (1) ☐ No (0)

Exclude if pregnant

**1.4. Patient transferred from another hospital?** ☐ Yes (1) ☐ No (0)

Exclude if transferred from another hospital

**1.5. Presentation to ED mostly due to infection?** ☐ Yes (1) ☐ No (0)

Exclude if presentation to ER was not due to infection

- **What is the initial Dx in ER?** ____________________________________________________

**1.6. Patient will be admitted to the hospital?** ☐ Yes (1) ☐ No (0)

Exclude if patient discharged from ED

**1.7. Patient admitted to the hospital through ED?** ☐ Yes (1) ☐ No (0)

Exclude if patient electively admitted to the hospital

**1.8. Code status of patient is "Do-Not-Resuscitate" (DNR)?** ☐ Yes (1) ☐ No (0)

Exclude if patient's code status is "DNR"

**1.9. Informed consent obtained?** ☐ Yes (1) ☐ No (0)

Exclude if patient refused to participate in the study (ONLY if recruiting center required informed consent)

### Enrolment

#### Demographics

**2.1.1. Inclusion date:** __ /__ /____ (DD/MM/YYYY)

**2.1.2. Race**

- ☐ Asian (1)
- ☐ Black (2)
- ☐ South Asian (3)
- ☐ White (4)

### History

**2.2.1. Patient known case of chronic respiratory diseases (e.g., asthma, COPD, ILD)?** ☐ Yes (1) ☐ No (0)

**2.2.2. Patient known case of myocardial infarction?** ☐ Yes (1) ☐ No (0)

**2.2.3. Patient known case of dementia?** ☐ Yes (1) ☐ No (0)

**2.2.4. Patient known case of heart failure?** ☐ Yes (1) ☐ No (0)

**2.2.5. Patient known case of peripheral vascular disease?** ☐ Yes (1) ☐ No (0)

**2.2.6. Patient known case of stroke or TIA?** ☐ Yes (1) ☐ No (0)

**2.2.7. Patient known case of rheumatological or autoimmune diseases?** ☐ Yes (1) ☐ No (0)

**2.2.8. Patient known case of peptic ulcer disease?** ☐ Yes (1) ☐ No (0)

**2.2.9. Patient known case of liver disease?**
- ☐ No (0)
- ☐ Mild (1)
- ☐ Moderate to severe (2)

**2.2.10. Patient known case of diabetes mellitus?**
- ☐ None (0)
- ☐ Diet-controlled (1)
- ☐ Uncomplicated (2)
- ☐ End-organ damage (3)

**2.2.11. Patient known case of hypertension or using hypertension medications?** ☐ Yes (1) ☐ No (0)

**2.2.12. History of smoking?** ☐ Yes (1) ☐ No (0)

**2.2.13. Patient known case of hemiplegia/paraplegia?** ☐ Yes (1) ☐ No (0)

**2.2.14. Patient known case of chronic kidney disease (CKD)?** ☐ Yes (1) ☐ No (0)

**2.2.15. Patient known case of cancer (solid tumor)?**
- ☐ No (0)
- ☐ Localized (1)
- ☐ Metastatic (2)

**2.2.16. Patient known case of leukemia?** ☐ Yes (1) ☐ No (0)

**2.2.17. Patient known case of lymphoma?** ☐ Yes (1) ☐ No (0)

**2.2.18. Patient known case of AIDS?** ☐ Yes (1) ☐ No (0)

#### Vital signs and blood tests

**2.3.1. Altered mental status (GCS <15)?** ☐ Yes (1) ☐ No (0)

**2.3.2. AVPU Score?**
- ☐ Alert (0)
- ☐ Reacts to voice (1)
- ☐ Reacts to pain (2)
- ☐ Unresponsive (3)

**2.3.3. Hypercapnic respiratory failure ($PaCO_2$ >45 mmHg ± pH <7.35)?** ☐ Yes (1) ☐ No (0)

**2.3.4. Body temperature: ______ °C**

**2.3.5. Heart rate: ______ beats/minute**

**2.3.6. Systolic blood pressure (SBP): ______ mmHg**

**2.3.7. Diastolic blood pressure (DBP): ______ mmHg**

**2.3.8. Respiratory rate: ______ breaths/minute**

**2.3.9. Oxygen saturation: ______ %**

**2.3.10. Any supplemental oxygen?** ☐ Yes (1) ☐ No (0)

**2.3.11. White blood cell (WBC) count: ______ $\times 10^3/\mu L$**

### In-hospital follow-up (30-days)

#### Infection status

**3.1.1. Was the patient discontinued from the study?** ☐ Yes (1) ☐ No (0)

**If yes to question 3.1.1.:**
- **Date of discontinuation:** __ / __ / ____ (DD/MM/YYYY)
- **Reason(s) for discontinuation:** ______________________________

**3.1.2. Was the initial presentation to ED due to infection?** ☐ Yes (1) ☐ No (0)

Exclude if initial presentation was not due to infectious cause

**3.1.3. If yes, infection confirmed by:**
- ☐ Positive body fluid(s) culture (1)
- ☐ Other microbiological methods (e.g., serology, PCR, … etc.) (2)
- ☐ Radiological evidence (3)
- ☐ Clinical context (4)

**3.1.4. If yes to 3.1.2.: What was the source of infection?**
- ☐ COVID-19 (1)
- ☐ Respiratory (except COVID-19) (2)

- ☐ Urinary (3)
- ☐ Abdominal (4)
- ☐ Soft tissue/cutaneous (5)
- ☐ Neurological (6)
- ☐ Bone and joints (7)
- ☐ Bloodstream (8)
- ☐ Unknown (9)
- ☐ Other (10), specify please: ___________________________________________________

#### In-hospital mortality (30-days)

**3.2.1. What is the status of patient by end of in-hospital follow-up?**

- ☐ Died (1)
- ☐ Discharged from hospital (2)
- ☐ Transferred to another hospital (3)
- ☐ Still hospitalized (4)

**3.2.2. If died, what was the cause of death?**

- ☐ COVID-19 (1)
- ☐ Heart ds (MI, CHF, dysrhythmia, ... etc.) (2)
- ☐ Respiratory ds (COPD, BA, respiratory failure, ... etc.) (3)
- ☐ Sepsis-related (septic shock, MODS, ... etc.) (4)
- ☐ Cancer (Solid or hematological) (5)
- ☐ Stroke (including intracranial hemorrhage) (6)
- ☐ Other (7), specify please: ___________________________________________________

**3.2.3. If died, specify date of death:** __ / __ / ____ (DD/MM/YYYY)

**3.2.4. If discharged from hospital, specify date of discharge:** __ / __ / ____ (DD/MM/YYYY)

**3.2.5. If transferred to another hospital, specify date of transfer:** __ / __ / ____ (DD/MM/YYYY)

#### ICU-admission (30-days)

**3.3.1. Has patient admitted to ICU?** ☐ Yes (1) ☐ No (0)

- **If admitted to ICU, specify date of ICU admission:** __ / __ / ____ (DD/MM/YYYY)

**3.3.2. Has patient discharged from ICU?** ☐ Yes (1) ☐ No (0)

- **If discharged from ICU, specify date of discharge from ICU:** __ / __ / ____ (DD/MM/YYYY)

### Out-hospital follow-up (90-days)

#### All-cause mortality (90-days)

**4.1. What is the status of patient by end of in-hospital follow-up?**

- ☐ Died (1)
- ☐ Alive (2)

**4.2. If died, what was the cause of death?**

- ☐ COVID-19 (1)

☐ Heart ds (MI, CHF, dysrhythmia, ... etc.) (2)
☐ Respiratory ds (COPD, BA, respiratory failure, ... etc.) (3)
☐ Sepsis-related (septic shock, MODS, ... etc.) (4)
☐ Cancer (Solid or hematological) (5)
☐ Stroke (including intracranial hemorrhage) (6)
☐ Other (7); specify please: ______________________________________________

**4.3. If died, specify date of death:** __ / __ / ____ (DD/MM/YYYY)
